# Effectiveness of ChAdOx1 nCoV-19 Corona Virus Vaccine (Covishield™) in preventing SARS-CoV2 infection, Chennai, Tamil Nadu, India, 2021

**DOI:** 10.1101/2022.04.15.22273859

**Authors:** Sharan Murali, Manikandanesan Sakthivel, Kamaraj P, Vettrichelvan Venkataswamy, Jeromie Wesley Vivian Thangaraj, Anita Shete, Alby John, Jaganathan Arjun, Girish Kumar C P, Pragya D Yadav, Rima Sahay, Triparna Majumdar, Manisha Dhudhmal, Azhagendran Sivalingam, Sudha Rani D, Augustine D, Vijayaprabha R, Murali Mohan M, Suresh A, Punita M, Elavarasu G, Prabhakaran C, Dhana Priya Vadhani S, Prakash M, Ezhil P, Ganeshkumar Parasuraman, Jagadeesan M, Manish S Narnaware, Gagandeep Singh Bedi, Prabhdeep Kaur, Manoj V Murhekar

## Abstract

**Background:** India experienced the second wave of the COVID-19 pandemic in March 2021, driven by the delta variant. Apprehensions around the usefulness of vaccines against delta variant posed a risk to the vaccination program. Therefore, we estimated the effectiveness of two doses of the ChAdOx1 nCoV-19 (Covishield) vaccine against COVID-19 infection among individuals ≥45 years in Chennai, India.

**Methods:** A community-based cohort study was conducted from May to September 2021 in a selected geographic area in Chennai, Tamil Nadu. The estimated sample size was 10,232. We enumerated individuals from all eligible households and periodically updated vaccination and COVID-19 infection data. We computed vaccine effectiveness with its 95% confidence interval for two doses of the Covishield vaccine against any COVID-19 infection.

**Results:** We enrolled 69,435 individuals, of which 21,793 were above 45 years. Two dose coverage of Covishield in the 18+ and 45+ age group was 18% and 31%, respectively. The overall incidence of COVID-19 infection was 1099 per 100,000 population. The vaccine effectiveness against COVID-19 disease in the ≥45 age group was 61.3% (95% CI: 43.6 - 73.4) at least two weeks after receiving the second dose of Covishield. Genomic analysis of 74 (28 with two doses, 15 with one dose, and 31 with zero dose) out of the 90 aliquots collected from the 303 COVID-19 positive individuals in the 45+ age group showed delta variants and their sub-lineages.

**Conclusion:** We demonstrated the effectiveness of two doses of the ChAdOx1 vaccine against the delta variant in the general population of Chennai. We recommend similar future studies considering emerging variants and newer vaccines. Two-dose vaccine coverage could be ensured to protect against COVID-19 infection.

## Introduction

World Health Organization (WHO) declared COVID-19 disease as a pandemic in March 2020 (1). India reported its first COVID-19 case on January 30, 2020 (2). The first wave of the disease in India peaked in mid-September and declined gradually by October 2020. The country witnessed the second wave from February to July 2021, with new cases reaching over 400,000 per day (3). COVID-19 deaths were reportedly higher among older adults and individuals with chronic morbid conditions (2). Chennai, the capital city of Tamil Nadu state in India, was one of the hotspots. Delta variant of the SARS-CoV2 virus drove the second wave, which peaked in early May 2021 in Chennai, India.

The Government of India (GoI) approved two COVID-19 vaccines for public use in January 2021, namely Covishield and Covaxin (4). Covishield is the brand name for the ChAdOx1 nCoV-19 Corona Virus Vaccine (Recombinant) manufactured by the Serum Institute, Pune, India. ChAdOx1 is a chimpanzee adenovirus – vector vaccine with SARS-CoV2 Spike protein, similar to the AZD1222 COVID-19 vaccine, manufactured by AstraZeneca (5). The second vaccine, Covaxin (BBV152), was the whole virion inactivated SARS-CoV2 vaccine (BBV152) manufactured by Bharat Biotech in association with the Indian Council of Medical Research (ICMR) - National Institute of Virology (NIV) (6). The Greater Chennai Corporation (GCC), the governing body of Chennai city, Tamil Nadu, India, included both vaccines in the COVID-19 vaccination drive.

India started COVID-19 vaccination on March 1, 2021, for individuals aged 60+ years and individuals with comorbidities in the 45+ age group. The Government expanded the vaccine program in a phased manner to different age groups depending on the availability of vaccines and the preparedness of the health system. From April 1, 2021, India expanded vaccination to all above 45 years and further added 18-45 years from May 1, 2021, onwards.

The second wave’s initiation coincided with the vaccination program’s expansion to the general population in March 2021. The majority of the COVID-19 infections (70%) in Tamil Nadu, India, between December to May 2021 were due to delta variant (8). Pooled analysis from three randomized controlled trials conducted in the United Kingdom, Brazil, and South Africa documented the efficacy of ChAdOx1 nCoV-19 (AZD1222) at around 70% (7). Few studies demonstrated the immune evasion property of the delta variant (B.1.617.2) and reduced effectiveness of vaccines (9–11). Although there is ample evidence for the effectiveness of the COVID-19 vaccine in preventing severe diseases (12–14), there was limited data regarding its effectiveness in preventing the infection.

In this context, a collaborative team from GCC and a public health research institution conducted a community-based study documenting vaccine effectiveness (VE). Our primary objective was to estimate the effectiveness of two doses of the ChAdOx1 nCoV-19 (Covishield) vaccine against RT-PCR confirmed COVID-19 infection among the individuals ≥45 years during the second wave of COVID-19 driven by the delta variant, Chennai, India. We also determined the distribution of SARS-CoV-2 Variants of concern (VoC) among a sample of individuals diagnosed with COVID-19.

## Methods

### Study design and setting

We conducted a community-based cohort study in Chennai from May 22, 2021, to September 30, 2021. Chennai is India’s 4th largest metropolitan city and the capital city of Tamil Nadu State, India. Greater Chennai Corporation (GCC) is divided into north, south, and central regions, with five zones each. Each zone is further divided into divisions, and there are 200 divisions. We established the cohort in three divisions and collected data using the COVID-19 surveillance system established by GCC (15).

Chennai registered its first COVID-19 cases on March 14, 2020. The city reported f 5,66,147 individuals positive for COVID-19 from the pandemic’s start till December 31, 2021. As a part of COVID-19 mitigation strategies, GCC has been conducting active surveillance, contact tracing, mobile testing, and positive patient follow-up. For active door-to-door, syndromic surveillance, GCC recruited temporary paid volunteers named Fever Survey Workers (FSWs) who visited every household at least twice a week to enquire about symptoms suggestive of COVID-19. They motivated individuals with at least one of these symptoms to get tested for COVID-19 free of cost at the nearest testing facility. In addition, symptomatic patients also sought care at the government or private health facilities. All health facilities across the city sent their samples for the RT-PCR tests for COVID-19 disease to the government-authorized laboratory. All laboratories uploaded the COVID-19 RT-PCR test results into a specially designed web portal named “*igotit*,” which generated line lists for every division.

FSWs collected the positive line list from the division office and contacted the patients. The division-level medical team triaged the positive individuals using a standardized protocol (Supplementary Figure 1). The FSWs also coordinated contact tracing and their testing. The medical team provided essential medication and counseling for patients in home isolation. The medical team referred the patients requiring hospitalization to designated COVID-19 hospitals based on the triaging. The telemedicine team at the zonal level monitored the patients for any red flags daily and directed them to the hospital if needed.

### Study Population and period

We conducted the study during the declining phase of the second wave of COVID-19 in Chennai City. The vaccination coverage among the eligible population (18+) was picking up rapidly (Supplementary figure 2). By May 22, 2021, about 20% of individuals ≥45 years received both doses of the COVID-19 vaccine.

#### Inclusion Criteria

We included all individuals who gave consent in the study area. India opened the COVID-19 vaccination camp for the 18-45 age group from May 1, 2021. Hence the coverage for this age group was low and limited to one dose only. Similarly, the coverage for Covaxin was only 12% in the study population. Therefore, we included only the 45+ age group for estimating the vaccine effectiveness of Covishield.

#### Exclusion Criteria

We excluded non-household settlements such as hostels, paying guests facilities, and residential hotels. All individuals who tested positive for COVID-19 during the study period from June 2021 to September 2021 were followed up to assess their disease outcomes.

### Sample Size & Sampling Strategy

Given the high transmission of variants of concern, we assumed 70% VE for the sample size calculation (7,16,17). Based on the surveillance data for the two peak months in the first wave, we considered the attack rate among the unvaccinated group as 2%, the confidence level of 95%, power of 80% and desired precision width of 15%. The minimum required sample size was 13,467 individuals ≥45 years (18).

We selected three geographically contiguous divisions - Divisions 147, 151 & 153, considering feasibility, representation of various socioeconomic groups, and rising trends in cases. In each selected division, we enumerated all the households which satisfied the eligibility criteria. The enumerators listed all the individuals in the selected households.

## Data Collection

We formed field teams consisting of supervisors with research experience and GCC Fever Survey Workers. The FSWs visited each household in the area assigned to them to assess their eligibility. After obtaining written informed consent from the eligible households, we assigned a unique identification number. We collected baseline data, including socio-demographic details, residential area details (slum/non-slum), and housing patterns (number of living rooms & total individuals in the house). We collected comorbidity status, history of previous COVID-19 infection, current COVID-19 disease, and vaccination details using an Open Data Kit Tool (ODK) (19). We updated the exposure and outcome data periodically until September 30, 2021.

### Exposure Assessment

We confirmed the vaccination status by verifying the vaccination card given to the patient (based either on the message received in the beneficiary phone number or the vaccination certificate downloaded from the portal exclusively maintained by MoHFW for COVID-19 vaccination (COWIN). We defined unvaccinated individuals as those who have not received the COVID-19 vaccine or were within 14 days of receiving the first dose.

We defined vaccination with single-dose as individuals who had completed 14 days after receiving the first dose of vaccine and had not received the second dose. The individuals who received the second dose but were within 14 days were also considered in this category. Vaccination with two doses included individuals who had completed 14 days after receiving the second dose of the vaccine.

## Outcome Measures

The primary outcome measure was RT-PCR confirmed COVID-19 infection. We collected the line list of positive individuals. Fourteen days after the confirmation of diagnosis, the data collectors telephonically interviewed the individuals. We ascertained the clinical outcome of the COVID-19 infection as recovered or dead. Among recovered, we collected hospitalization details and documented the self-reported need for ventilator or oxygen support. If a COVID-19 positive individual died within 28 days from the date of diagnosis of the COVID-19 disease, we considered the event as COVID death. However, death due to non-natural causes (e.g., accidental, intentional self-harm, and homicide) was not deemed as COVID-19 death even if it was within 28 days from the date of diagnosis for COVID-19 disease (20).

## Sample collection and Laboratory Investigations

We collected nasal and oro-pharyngeal (N/OP) swabs from RT-PCR confirmed COVID-19 patients aged above 45 years. The team shipped an aliquot of the COVID-19 positive samples to ICMR-National Institute of Virology, Pune for Next-Generation Sequencing (NGS).

Viral RNA was extracted (N/OP) from swab samples using an RNA extraction kit (Applied Biosystem, Waltham, MA, USA) and tested for the detection of the E gene using Real-time RT-PCR)(21). We included samples having Ct <30 for NGS. In brief, the COVIDSeq protocol (Illumina Inc, USA) includes amplification, tagmentation of the cDNA, and quantification of purified libraries using the KAPA Library Quantification Kit (Kapa Biosystems, Roche Diagnostics Corporation, USA). For sequencing, pooled libraries were denatured, neutralized, and loaded at a concentration of 1.4 pM onto the NextSeq 500/550 system using High Output Kit v2.5 (75 Cycles)(22). Bcl files generated were converted to fastq using bcl2fastq2 Conversion Software v2.20. We performed reference-based mapping using CLC Genomics Workbench v.22.0 to retrieve the sequence of the SARS-CoV-2. We generated the phylogenetic tree using MEGA software version 10.

## Statistical Analysis

For the primary analysis of the vaccine for Covishield, we excluded all individuals who were less than 45 years or took vaccines other than Covishield. We also excluded the individuals who had taken the first dose of the vaccine within 14 days or reported past infection with COVID-19.

We calculated the mean (standard deviation) and median (Inter-quartile range) for age. After which, we calculated frequencies and proportions for socio-demographics for overall respondents and 45-98 years of age group. We summarized the vaccine-related and outcome-related variables for each age group of 0-17, 18-44, 45-59, and 60-98 years and gender as proportions.

Unadjusted Relative Risk (RR) and 95% Confidence Interval (95% CI) of RTPCR-positive individuals who received the Covishield vaccine for 45-59 years and 60-98 years were calculated. We calculated Unadjusted Vaccine Effectiveness VE = 1 – RR and 95% Confidence Interval.

Further, we adjusted the relative risk for the age group 45-59 years and 60-98 years using Mantel–Haenszel stratified analysis and estimated the Adjusted Relative Risk (ARR) and 95% Confidence Interval. We used the relationship 1-ARR to calculate the final adjusted Vaccine Effectiveness with its 95% Confidence Interval for two doses of Covishield against any RT-PCR positive infection. We calculated power considering a 95% confidence level for two doses of Covishield in preventing infections and for severe disease. The data team used STATA SE (version 17.0) software (StataCorp LLC, Texas, USA) was for statistical analysis (23).

## Human Participant Protection

We obtained approval from the Institutional Ethics committee and obtained required administrative approvals. We conducted extensive training sessions for the Fever Survey Workers to collect data while ensuring privacy and comfort of respondents. We obtained written informed consent from the participants before enrolling them in the cohort.

FSWs referred individuals requiring emotional support during the follow-up visits. We conducted the interviews adhering to the COVID-19 personal protection guidelines and maintained complete confidentiality and anonymity of the data. Trained Laboratory technicians collected the throat and nasal swabs wearing full PPE Kit and followed all aseptic precautions.

## Results

### Socio-demographic Characteristics

Of the 20,913 households in the three divisions in Chennai, India, 19,211 (92%) were enrolled in the cohort (Figure 1). A total of 69,435 individuals were residing in the enrolled households. One-fifth (n=3,466; 18%) of the households were in slum areas, and 95.5% were pucca houses (Supplementary Table 1). Of the 69,435 individuals, nearly half (n=34,500; 49.7%) were males, and 31.4% (n=21,793) belonged to the 45+ age group (Supplementary Table 2). Over half (n=35,152; 50.6%) had formal school education, and 39.4% (n=27,365) had a college education. About 9% (n=4,912) of the individuals had a history of Diabetes Mellitus.

**Figure 1:**
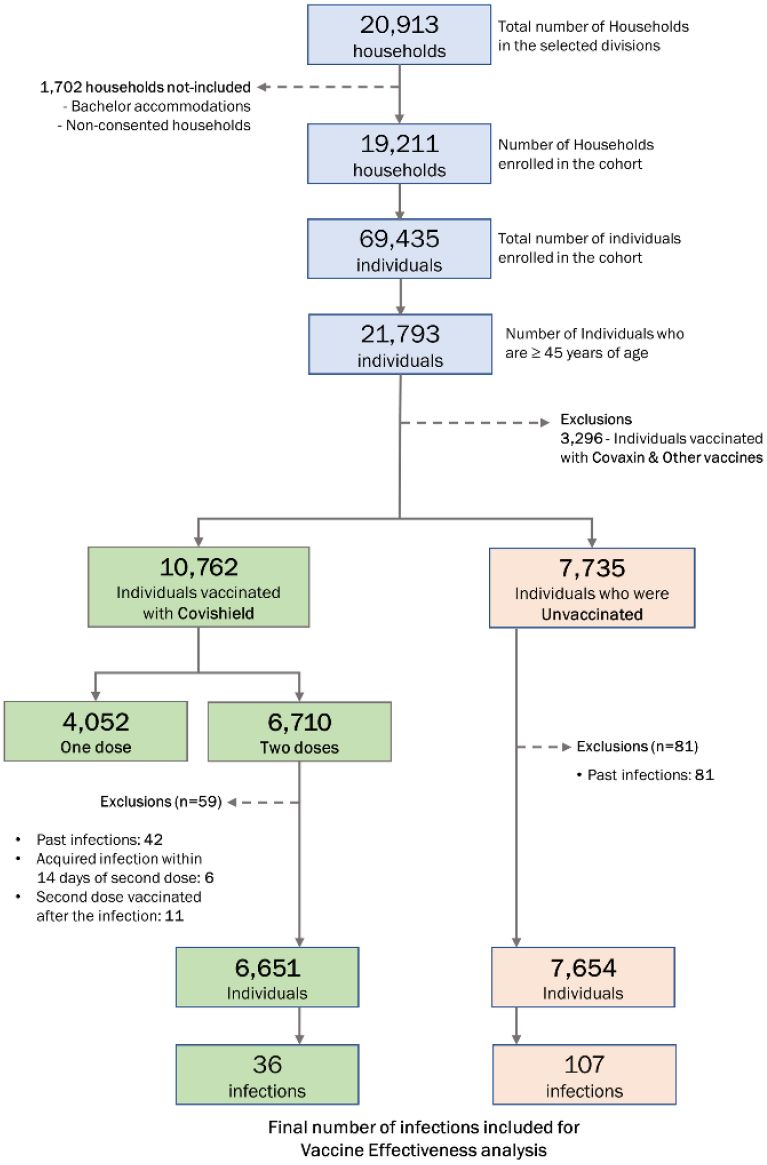
Participant enrollment, events, and data exclusions for estimating the effectiveness of ChAdOx1 nCoV-19 Corona Virus Vaccine (Covishield^™^) in reducing delta variant of SARS-CoV2 infection, Chennai, Tamil Nadu, India, 2021

We included 21,793 individuals ≥45 years for the analysis of vaccine effectiveness (Table 1). About 60% (n=13,114) belonged to the 45-59 age group, and nearly half (n=10,916; 50.1%) were males. The proportion of health care and frontline workers was 2.4% (n=532).

**Table 1.** Socio-demographic characteristics of the individuals ≥45 years included in the analysis of COVID-19 vaccine effectiveness cohort study, Chennai, 2021 (N=21,793)

| Characteristic |  | n | % |
| --- | --- | --- | --- |
| Division | 147 | 8382 | 38.5 |
|  | 151 | 4881 | 22.4 |
|  | 153 | 8530 | 39.1 |
| Gender | Male | 10,916 | 50.1 |
|  | Female | 10,844 | 49.8 |
|  | Transgender | 33 | 0.1 |
| Age group (in years) | 45 - 59 | 13,114 | 60.2 |
| | $\geq 60$ | 8,679 | 39.8 |
| Education | No formal education | 2,689 | 12.3 |
|  | School education | 11,749 | 54.0 |
|  | College education | 7,355 | 33.7 |
| Occupation | Health care / Frontline worker | 532 | 2.4 |
|  | Others | 21,261 | 97.6 |
| Comorbidity<br>(N=21788)* | Known HT | 3,770 | 17.3 |
|  | Known DM | 4,251 | 19.5 |
|  | Others | 904 | 4.1 |
| Vaccination status | Unvaccinated | 7,735 | 35.5 |
|  | Received only one dose | 4,972 | 22.8 |
|  | Received both the doses | 9,086 | 41.7 |
| Person per room | Person/room < 3 | 14,755 | 67.7 |
| | Person/room $\geq 3$ | 7,038 | 32.3 |
| Residential Area | Slum | 3,271 | 15.0 |
|  | Non Slum | 18,522 | 85.0 |

HT - Hypertension
DM - Diabetes Mellitus
\*Missing for five individuals

### Vaccination status

Of the 21,793 individuals in the 45+ age cohort, 7,735 (36%) were unvaccinated. About half (n=10,762; 49.4%) of the 45+ cohort population received Covishield vaccine (Table 2). The vaccination coverage was almost similar among males (n=7,191; 66%) and females (n=6,867; 63%). The coverage with at least one dose was highest (n=5,703; 65.7%) among the elderly (≥60 years), followed by the 45-59 age group (n=8,355; 63.7%).

**Table 2.**
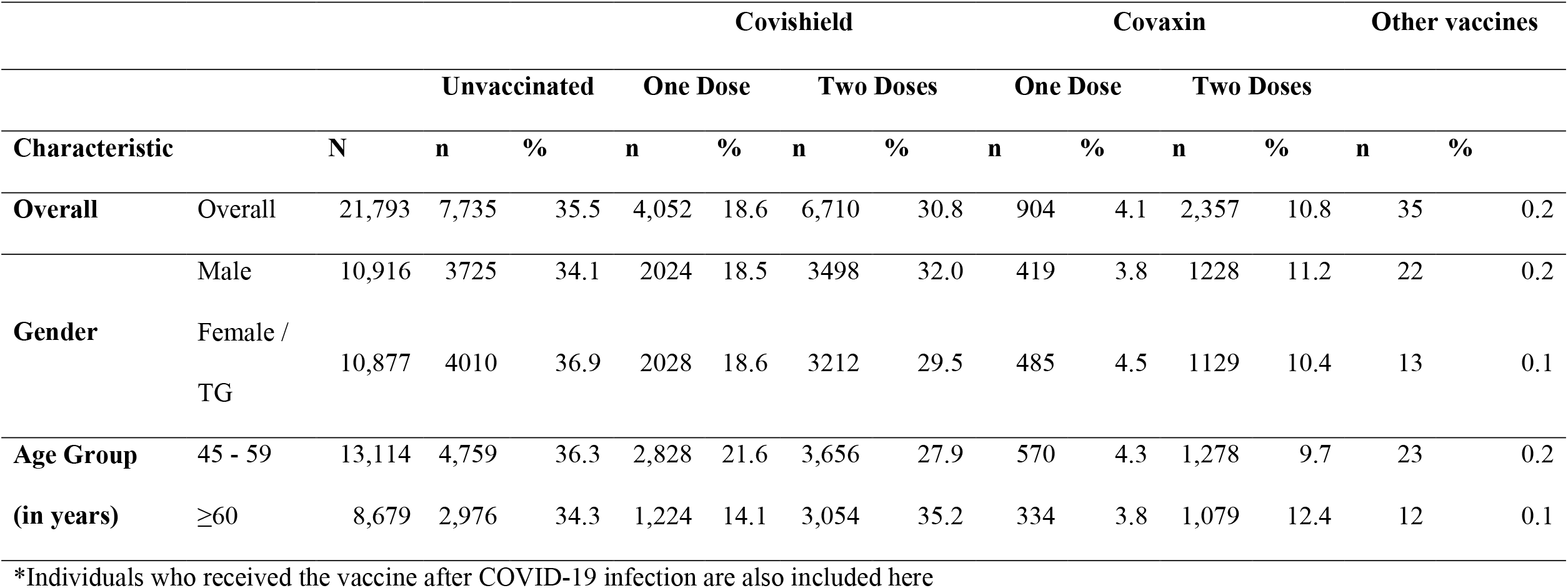
Vaccination profile of the individuals ≥45 years in the COVID-19 vaccine effectiveness cohort study, Chennai, 2021 (N=21,793)*

### COVID-19 disease outcomes

In the 45+ cohort of 21,793 individuals, 303 tested positive for COVID-19, with an incidence of 1,390 per 100,000 population during the study period (Supplementary Table 5). Among the 10,762 individuals ≥45 years included in the primary analysis of VE, 123 tested positive for COVID-19 during the study period (Table 3). The incidence was higher among females (1,260 per 100,000) than males (1,032 per 100,000). The incidence was almost similar in the 45-59 age group (1,141 per 100,000) and the 60+ age group (1,145 per 100,000).

**Table 3.**
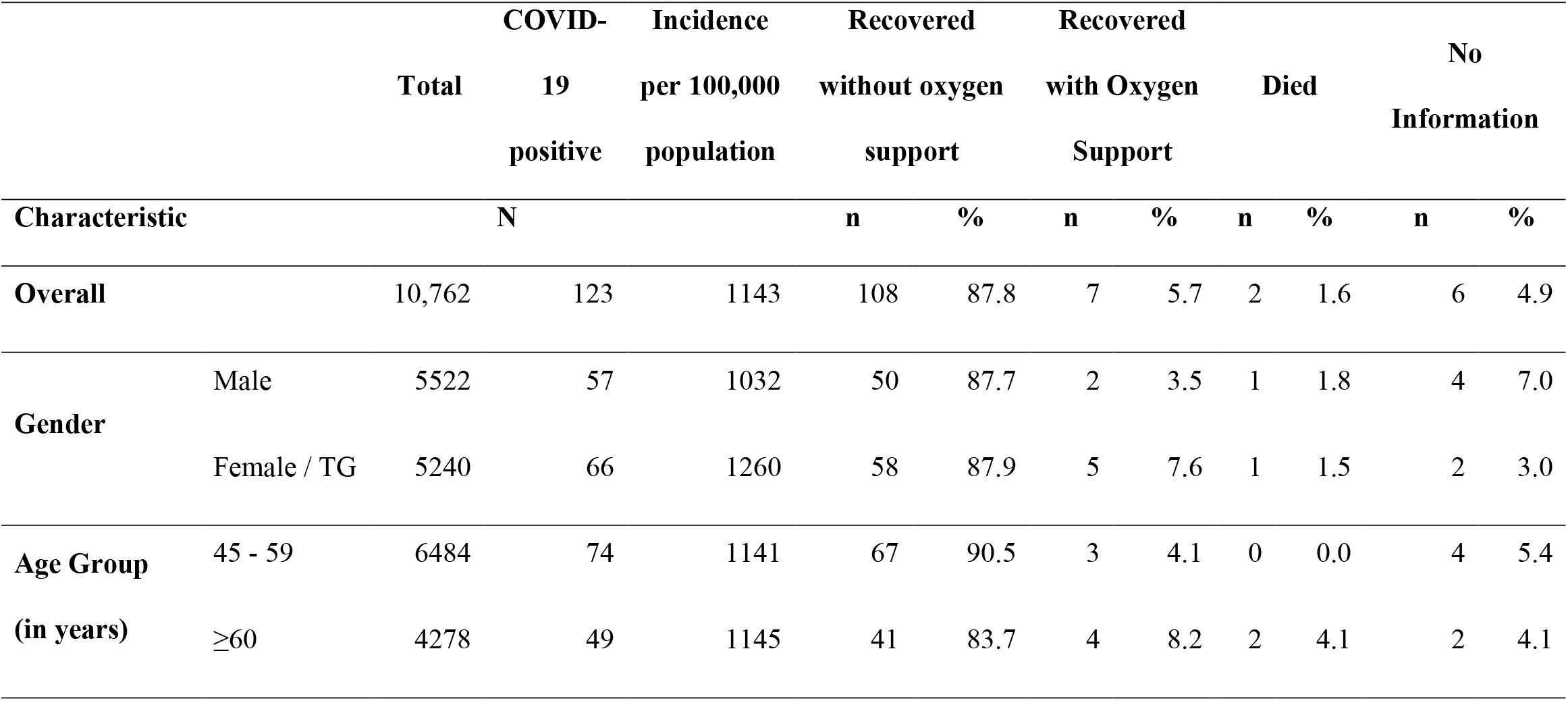
COVID-19 incidence and its outcomes among the individuals ≥45 years in the COVID-19 vaccine effectiveness cohort study, Chennai, 2021 (N=10,762)

Among the 123 individuals who had COVID-19, 5.7% (n=7) recovered with oxygen, and 87.8% (n=108) recovered without oxygen. The proportion of COVID-19 patients who required oxygen was 8.2% (n=4) in the 60+ age group and 4.1% (n=3) in the 45-59 age group (Table 3). Of the 2 (1.6%) COVID-19 deaths, both were ≥60 years of age.

### VE of two doses of Covishield

We excluded 3,296 individuals vaccinated with Covaxin and vaccines other than Covishield for the primary analysis of vaccine effectiveness (VE). Among the 7,735 unvaccinated individuals, we excluded 81 (1%) individuals who had past infections of COVID-19. Similarly, we excluded 59 (<1%) individuals from among the 6,710 individuals vaccinated with two doses of Covishield Vaccine from the analysis for VE analysis (Figure 1).

The overall relative risk (RR) against COVID-19 infection at least two weeks after receiving two doses of Covishield (ChAdOx1 nCoV-19) vaccine was estimated to be 0.37 (95% CI: 0.26-0.56). This translated to an overall vaccine effectiveness (VE) of 61.4% (95% CI: 43.6 – 73.4). The reported RR for the 45–59-year age group was 0.34 (95% CI: 0.20-0.58) and that for the 60+ age group was 0.45 (95% CI: 0.26-0.78). The estimated VE was higher among 45-59 years age group [65.6% (95% C.I: 42.3 - 79.5)] than 60+ age group [55.1% (95% C.I: 21.7 - 74.2)].

We also calculated the RR and VE for the various subgroups within the 45+ cohort. After adjusting for the number of individuals per room in the household, the VE estimates showed a maximum of 63.8% (95% CI: 46.8-75.3). The covariate-adjusted VE estimates looked almost closer to the overall VE reported in the study (Table 4).

**Table 4.** Effectiveness of two doses of Covishield vaccine against COVID-19 infection among individuals ≥45 years, Chennai, India (N=14,305)

| Characteristics & its category |  | Vaccination status | Total | COVID-19 positive |  | RR<br>(95% CI) | VE<br>(95% CI) | Adjusted RR*<br>(95% CI) | Adjusted VE*<br>(95% CI) |
| --- | --- | --- | --- | --- | --- | --- | --- | --- | --- |
|  |  |  | N | n | % |  |  |  |  |
| Overall | ≥45 years | Received two doses | 6651 | 36 | 0.5 | 0.386 (0.264 - 0.564) | 61.4 (43.6 - 73.6) |  |  |
|  |  | Unvaccinated | 7654 | 107 | 1.4 |  |  |  |  |
| Age Group | 45-59 years | Received two doses | 3628 | 18 | 0.5 | 0.344 (0.205 - 0.577) | 65.6 (42.3 - 79.5) | 0.386 (0.264 - 0.564) | 61.4 (43.6 - 73.6) |
|  |  | Unvaccinated | 4713 | 68 | 1.4 |  |  |  |  |
|  | ≥60 years | Received two doses | 3023 | 18 | 0.6 | 0.449 (0.258 - 0.783) | 55.1 (21.7 - 74.2) |  |  |
|  |  | Unvaccinated | 2941 | 39 | 1.3 |  |  |  |  |
| Gender | Male | Received two doses | 3463 | 18 | 0.5 | 0.376 (0.220 - 0.642) | 62.4 (35.8 - 78.0) | 0.388 (0.266 - 0.565) | 61.2 (43.5 - 73.4) |
|  |  | Unvaccinated | 3687 | 51 | 1.4 |  |  |  |  |
|  | Female / TG | Received two doses | 3188 | 18 | 0.6 | 0.400 (0.236 - 0.679) | 60.0 (32.1 - 76.4) |  |  |
|  |  | Unvaccinated | 3967 | 56 | 1.4 |  |  |  |  |
| Comorbidity | HT/DM | Received two doses | 1987 | 12 | 0.6 | 0.459 (0.231 - 0.916) | 54.1 (8.4 - 77.0) | 0.389 (0.266 - 0.567) | 61.1 (43.3 - 73.4) |
|  |  | Unvaccinated | 1826 | 24 | 1.3 |  |  |  |  |
|  | Non-HT/DM | Received two doses | 4663 | 24 | 0.5 | 0.361 (0.230 - 0.568) | 63.9 (43.2 - 77.0) |  |  |
|  |  | Unvaccinated | 5828 | 83 | 1.4 |  |  |  |  |
| Person per room | < 3 | Received two doses | 5034 | 31 | 0.6 | 0.389 (0.256 - 0.591) | 61.1 (40.9 - 74.4) | 0.362 (0.247 - 0.532) | 63.8 (46.8 - 75.3) |
|  |  | Unvaccinated | 4610 | 73 | 1.6 |  |  |  |  |
|  | ≥ 3 | Received two doses | 1617 | 5 | 0.3 | 0.277 (0.108 - 0.706) | 72.3 (29.4 - 89.2) |  |  |
|  |  | Unvaccinated | 3044 | 34 | 1.1 |  |  |  |  |
| Residential Area | Slum | Received two doses | 528 | 0 | 0.0 | 0.091 (0.005 - 1.512) | 90.9 (-51.2 - 99.5) | 0.369 (0.251 - 0.541) | 63.1 (45.9 - 74.9) |
|  |  | Unvaccinated | 1683 | 18 | 1.1 |  |  |  |  |
|  | Non-Slum | Received two doses | 6123 | 36 | 0.6 | 0.394 (0.268 - 0.580) | 60.6 (42.0 - 73.2) |  |  |
|  |  | Unvaccinated | 5971 | 89 | 1.5 |  |  |  |  |

1 VE - Vaccine effectiveness | C.I - Confidence interval | RR - Relative Risk
- 2 Individuals who were unvaccinated and vaccinated with two doses of Covishield only were included in this analysis (n=14,445).<sup>123</sup> individuals who reported past infection, Six individuals who acquired infection within 14 days of the second dose of Covishield and 11 who got vaccinated with second dose after the infection were excluded (n=140).
\* Mantel–Haenszel stratified adjusted relative risk and vaccine effectiveness

The power of the VE estimation for two doses of Covishield against any COVID-19 infection was 99.9%. However, the power for the VE estimate for two doses of Covishield against the severe disease was only 34%. The estimated VE for a single dose of Covishield against COVID-19 infection in the 45+ age group was 28.7% (95% CI: -2.3 - -50.3; power - 45%).

### Genomic sequencing results

We could collect aliquots for 90 samples out of the 303 COVID-19 patients in the >=45 years of age from the respective testing laboratory. Out of 90 samples received at ICMR NIV Pune, only 75 (83%) samples had a CT value less than 30. We processed these 30 samples for NGS. E gene copy numbers were found to be similar in the unvaccinated (7.7 × 10 1 to 2.1 ×108), one dose (5.2 × 10 1 to 6.0 × 108) and two-dose (7.1 × 10 1 to 3.4 × 109) vaccinated groups.

We could retrieve genome > 98% from 67 samples. Therefore, we performed GSAID on these 67 samples only. All the 67 samples tested positive for delta variants and sub-lineages, including AY. 100 (n=16), B.1.617.2 (n=14), AY.127 (n=11), AY.25.1 (n=8), AY.5.4 (n=4) AY. 50(n=3), AY. 122 (n=2), AY.59 (n=2) and one each with AY.11, AY.16, AY.126, AY.20, AY.54, and AY. 107, AY.86. We generated a Neighbours joining tree using MEGA version 10 with the Tamura-3-parameter model with bootstrap replication of 1000 cycles (Supplementary Figure 3).

Variant analysis in our study indicated the presence of E484Q mutation in 2 out of 67 delta sub-lineages belonging to AY.100 (EPI_ISL_9320575_V2) and AY.25.1 (EPI_ISL_9354202_V2), respectively. Both the patients were vaccinated with two doses of the Covishield vaccine. Two mutations of concern and interest in the RBD with evidence of increasing transmissibility or virulence, L452R, and T478K, were detected in 57 and 59 out of 67 samples, respectively. One of the mutations of concern in the furin cleavage site, P681R, was found in all the sequences retrieved in the current study. All the above mutations were seen as a result of transversion of single nucleotide variants (SNV) (Supplementary Figure 4).

## Discussion

We conducted this study towards the latter half of the second wave, which overwhelmed the health system due to increased hospitalization. The study was initiated to generate locally relevant evidence to build trust in the effectiveness of the COVID-19 vaccine. Our study documented vaccine effectiveness (VE) in a community cohort during the surge due to delta variant in a metropolitan city in India. Two doses of Covishield effectively protected the population above 45 years from COVID-19 infection. The study included all sections of a population, hence generating data closer to the real-world setting. It was challenging to develop a community cohort in a pandemic setting due to the high burden on the health care workers on the field, apprehensions among the population, and poor linkages between disease and vaccination data. The available resources of GCC, trust among the public towards FSWs, and complementary expertise of the collaborators enabled the planning and execution of this study.

Our study reiterates the protective effect of vaccines against RT-PCR confirmed COVID-19 infection as reported in three other studies from India. Our study’s VE estimates against COVID-19 disease were lower than those among a cohort of healthcare workers and comparable to two other studies from India (12–14). The first study had unique characteristics, such as a large nationwide cohort that included relatively younger healthy HCWs and FLWs. The cohort size was 1.6 million with 82% two-dose coverage of any COVID-19 vaccine and reported VE of 94.9% during January - May 2021 (12). The second study conducted during April - May 2021 using a test-negative case-control design among 4360 individuals (2379 cases and 1981 controls) estimated 63% VE for two doses of Covishield in Delhi, India (14). The third study was among a cohort of 10,567 health care workers, predominantly vaccinated with Covishield, and reported a VE of 65% from Feb to May 2021 in Tamil Nadu, India (13). Irrespective of the study design or study population, the results consistently supported the usefulness of two doses of Covishield.

The VE in our study was also comparable to the studies from other countries for ChAdOx1 nCoV-19 (AZD1222) vaccine. The efficacy was 62% in a multi-centric trial across the UK, Brazil, and South Africa involving 11636 participants for two doses of AZD1222 during April - November 2020 (24). Subsequently, the surveillance data from the UK up to September 2021 documented 60-70% VE against infection (25). A meta-analysis of studies reporting VE against delta estimated a pooled VE of 64% (57-71) after two doses of AZD1222 (26). Given the evidence globally and from India, the vaccination program should focus on ensuring two-dose coverage of ChAdOx1 nCoV-19 (Covishield) to reduce the risk of COVID-19 infection.

Next-generation sequencing methods provide an added advantage in understanding the genetic epidemiology of the COVID-19 infected population. The present investigation revealed the predominance of delta and its sub lineages during the study period. Variant analysis supported the transmissibility of the circulating delta lineages in the current study.

India’s vaccination program had two approved vaccines produced in India during the study period. The availability of manufacturing facilities in the country enabled rapid scaling of the vaccination program. A metanalysis pooled available data for multiple vaccines against delta variant infection and estimated the VE to be 84% for BNT162b2 (Pfizer–BioNTech) compared to 64% for AZD1222 (ChAdOx1 nCoV-19). Despite lower VE of AZD1222 vaccine, availability at scale and operationalization of vaccination program offered reasonable protection to the population against delt variant infection.

One of the limitations of the present study was the failure to ascertain the previous infection in the cohort using serology. However, the proportion of asymptomatic infection might have been similar in both vaccinated and non-vaccinated groups. The second limitation was an underestimation of symptomatic infections. We tried to overcome this limitation by using active surveillance for symptomatic infections and testing all symptomatic individuals and contacts. We pooled the line list of all COVID-19 positive individuals tested in any of the labs in the city to ensure the completeness and representativeness. However, we might have missed individuals who did not reveal their symptom status and did not get tested.

The third limitation was the inadequate sample size to study the VE against severe infections. Our primary objective was to understand the protective effect of COVID-19 vaccines irrespective of the severity of disease in the general population. As the cases started declining after May 2021, the number of hospitalization were inadequate for subgroup analysis. However, there is adequate evidence to support the usefulness of COVID-19 vaccines in preventing severe infection in India. Two hospital-based case-control studies among RT-PCR tested individuals reported around 80% effectiveness of Covishield (14,27). The effectiveness was 92-94% among a cohort of HCWs predominantly vaccinated with Covishield (13).

## Conclusion

Two doses of the Covishield (ChAdOx1 nCoV-19) vaccine protected the adult population from the delta variant of COVID-19 infection in Chennai, India. We may require similar VE studies at the population level in the context of changing variants and newer vaccines and boosters. The disease surveillance and vaccination program databases are currently separate, limiting the opportunities for systematic periodic analysis of outcomes. We recommend linkages between the disease surveillance and vaccination databases at the district or state level to enable regular in-depth real-time analysis. High vaccination coverage of two doses should be ensured to maintain population-level immunity against COVID-19 infection.

## Data Availability

All data produced in the present study are available upon reasonable request to the authors

## Author Contributions

SM and MS contributed equally to the paper as joint first authors. MM, PK, PG, MS, and SM designed the study, planned data analysis, and wrote the first draft of the paper. KP and VV managed and analyzed the data and reviewed the manuscript. JT, GK, and SR helped in designing, and data acquisition, and gave critical comments on the genome sequencing component of the study. AS, PY, RS, TM, and MD did data acquisition, and prepared the first draft of the genome sequencing component of the study. AS, AD, VR, MM, SA, PM, PM, DS, PC, EP, and EG did data acquisition, and analysis. JA, JM, MN, and GB coordinated the data acquisition, interpreted the data, and helped revise the manuscript. JT, and MM gave critical comments and revised the manuscript. All authors approved the final manuscript. PK is the guarantor. The corresponding author attests that all listed authors meet authorship criteria and that no others meeting the criteria have been omitted. The views presented here are those of the authors and should not be attributed to ICMR-NIE.

## Acknowledgements

We thank all the Greater Chennai Corporation health staff, including Zonal Health Officers, doctors, nurses, field staff, volunteers who coordinated with the research team in door-to-door data collection. We extend our sincere gratitude to the printing press department of Greater Chennai Corporation for providing the required prints in a timely manner. We acknowledge the contribution of Aswini S, Amirthammal Guna Grace, Vignesh, G Keerthiga, and Swapna R who provided the required technical support to the data team of the study. We thank the technical staff R Sudha, P. Lourdu Stella Mary, M.R. Santhi, I. Kalaimani, T. Ravichandran, T. Subba Rao, K. Ramu, P. Shantha, and Gandhimathi who helped in physical scrutiny of the data entry forms being filled from the field. We also thank the “*igotit*” data team who provided the required support in their portal to accommodate the vaccine coverage data. Authors are grateful for the excellent support provided by Dr. Abinendra Kumar, Mrs. Savita Patil, Mr. Yash Joshi, Ms Pranita Gawande, Ms. Jyoti Yemul for processing of the samples for NGS.

