## Supplementary files for "Effectiveness of ChAdOx1 nCoV-19 Corona Virus Vaccine (Covishield™) in preventing SARS-CoV2 infection, Chennai, Tamil Nadu, India, 2021"

**SUPPLEMENTARY FILE**

| **TABLES** | |
| --- | --- |
| 1 | Socio-demographic characteristics of the households participated in the COVID-19 vaccine effectiveness cohort study, Chennai, 2021 (N=19211) |
| 2 | Socio-demographic characteristics of the individuals who participated in the COVID-19 vaccine effectiveness cohort study, Chennai, 2021 (N=69435) |
| 3 | Supplementary Table 3. Vaccination profile of the vaccinated individuals in the COVID-19 vaccine effectiveness cohort study, Chennai, 2021 (N=30543) |
| 4 | COVID-19 incidence and its outcomes among the individuals in the  COVID-19 vaccine effectiveness cohort study, Chennai, 2021 (N=69,435) |
| 5 | COVID-19 incidence and its outcomes among individuals ≥45 years in the  COVID-19 vaccine effectiveness cohort study, Chennai, 2021 (N=21,793) |
| 6 | Effectiveness of one dose of Covishield vaccine against COVID-19 infection among individuals ≥45 years, Chennai, India (N=11,668) |
| **FIGURES** | |
| 1 | Tamil Nadu COVID-19 Case Management Protocol |
| 2 | Vaccine coverage among the eligible population (18+) and the total number of reported COVID-19 cases by week, Chennai, Tamil Nadu, India, January – December 2021 |
| 3 | Neighbour-joining tree of the SARS-CoV-2 sequences retrieved from the clinical samples collected from the 45+ age group individuals who were positive for SARS-CoV-2 in COVID-19 Vaccine effectiveness Study, Chennai, India, 2021. |
| 4 | Single nucleotide variations in the SARS CoV-2 sequences retrieved from the clinical samples collected from the 45+ age group individuals who were positive for SARS-CoV-2 in COVID-19 Vaccine effectiveness Study, Chennai, India, 2021. |

| **Supplementary Table 1. Socio-demographic characteristics of the households participated in the COVID-19 vaccine effectiveness cohort study, Chennai, 2021 (N=19211)** | | | |
| --- | --- | --- | --- |
| **Characteristic** |  | **n** | **%** |
| Division | 147 | 7,555 | 39.3 |
|  | 151 | 4,155 | 21.6 |
|  | 153 | 7,501 | 39.1 |
| Area | Slum | 3,466 | 18.0 |
|  | Non-Slum | 15,745 | 82.0 |
| Type of house | Kucha | 197 | 1.0 |
|  | Semi pucca | 658 | 3.4 |
|  | Pucca | 18,356 | 95.5 |
| Ration card (indicative of socio-economic status) | Below poverty line (BPL) | 1,101 | 5.7 |
|  | Non-BPL | 15,086 | 78.5 |
|  | No card | 3,024 | 15.7 |
| Number of family members | <4 | 8,831 | 46.0 |
|  | ≥4 | 10,380 | 54.0 |
| Rooms for sleeping | 1 | 6,312 | 32.9 |
|  | 2 | 10,113 | 52.6 |
|  | ≥3 | 2,786 | 14.5 |
| Number of people per room | 1 | 2,765 | 14.4 |
|  | 2 | 10,324 | 53.7 |
|  | 3 | 4,069 | 21.2 |
|  | ≥4 | 2,053 | 10.7 |

| **Supplementary Table 2. Socio-demographic characteristics of the individuals who participated in the COVID-19 vaccine effectiveness cohort study, Chennai, 2021 (N=69435)** | | | |
| --- | --- | --- | --- |
| **Characteristic** |  | **n** | **%** |
| Division | 147 | 27760 | 40.0 |
|  | 151 | 15090 | 21.7 |
|  | 153 | 26585 | 38.3 |
| Gender | Male | 34,500 | 49.7 |
|  | Female | 34,836 | 50.2 |
|  | Transgender | 99 | 0.1 |
| Age group (in years) | 0 - 17 | 15,135 | 21.8 |
|  | 18 - 44 | 32,507 | 46.8 |
|  | 45 - 59 | 13,114 | 18.9 |
|  | 60 - 98 | 8,679 | 12.5 |
| Education | No formal education | 6,918 | 10.0 |
|  | School education | 35,152 | 50.6 |
|  | College education | 27,365 | 39.4 |
| Occupation | Health care / Frontline worker | 1,418 | 2.0 |
|  | Others | 68,017 | 98.0 |
| Comorbidity | Known HT | 4,283 | 7.9 |
|  | Known DM | 4,912 | 9.0 |
|  | Others | 1,138 | 2.1 |
| Vaccination status | Unvaccinated | 38,892 | 56.0 |
|  | Received only one dose | 16,700 | 24.1 |
|  | Received both the doses | 13,843 | 19.9 |
| HT - Hypertension |  |  |  |
| DM - Diabetes Mellitus |  |  |  |

| **Supplementary Table 3. Vaccination profile of the vaccinated individuals in the**  **COVID-19 vaccine effectiveness cohort study, Chennai, 2021 (N=30543)** | | | | | | |
| --- | --- | --- | --- | --- | --- | --- |
|  |  |  | **Single** | | **Double** | |
| **Characteristic** |  | **N** | **n** | **%** | **n** | **%** |
| **Overall** |  | 30543 | 16700 | 54.7 | 13843 | 45.3 |
| **Gender** | Male | 15342 | 8191 | 53.4 | 7151 | 46.6 |
|  | Female / TG | 15201 | 8509 | 56.0 | 6692 | 44.0 |
| **Age group (in years)** | 18 - 44 | 16485 | 11728 | 71.1 | 4757 | 28.9 |
|  | 45 - 59 | 8355 | 3411 | 40.8 | 4944 | 59.2 |
|  | ≥60 | 5703 | 1561 | 27.4 | 4142 | 72.6 |
| **Covishield (N=23578)** |  |  |  |  |  |  |
| **Overall** |  | 23578 | 13947 | 59.2 | 9631 | 40.8 |
| **Gender** | Male | 11932 | 6942 | 58.2 | 4990 | 41.8 |
|  | Female / TG | 11646 | 7005 | 60.1 | 4641 | 39.9 |
| **Age group (in years)** | 18 - 44 | 12816 | 9895 | 77.2 | 2921 | 22.8 |
|  | 45 - 59 | 6484 | 2828 | 43.6 | 3656 | 56.4 |
|  | ≥60 | 4278 | 1224 | 28.6 | 3054 | 71.4 |
| **Covaxin (N=6852)** |  |  |  |  |  |  |
| **Overall** |  | 6852 | 2684 | 39.2 | 4168 | 60.8 |
| **Gender** | Male | 3337 | 1207 | 36.2 | 2130 | 63.8 |
|  | Female / TG | 3515 | 1477 | 42.0 | 2038 | 58.0 |
| **Age group (in years)** | 18 - 44 | 3591 | 1780 | 49.6 | 1811 | 50.4 |
|  | 45 - 59 | 1848 | 570 | 30.8 | 1278 | 69.2 |
|  | ≥60 | 1413 | 334 | 23.6 | 1079 | 76.4 |
| *Individuals who received the vaccine after COVID-19 infection are also included here | | | | | | |

| **Supplementary Table 4. COVID-19 incidence and its outcomes among the individuals in the  COVID-19 vaccine effectiveness cohort study, Chennai, 2021 (N=69,435)** | | | | | | | | | | | | | | |
| --- | --- | --- | --- | --- | --- | --- | --- | --- | --- | --- | --- | --- | --- | --- |
|  |  | **Total** | **COVID-19 positive** | **Incidence per 100,000 population** | **Recovered without oxygen support** |  | **Recovered with Oxygen Support** |  | **Recovered with ventilator support** |  | **Died** |  | **No Information** |  |
|  |  |  | **N** |  | **n** | **%** | **n** | **%** | **n** | **%** | **n** | **%** | **n** | **%** |
| **Overall** |  | 69435 | 763 | 1099 | 701 | 91.9 | 24 | 3.1 | 2 | 0.3 | 16 | 2.1 | 20 | 2.6 |
| **Gender** | Male | 34500 | 365 | 1058 | 334 | 91.5 | 10 | 2.7 | 1 | 0.3 | 8 | 2.2 | 12 | 3.3 |
|  | Female / TG | 34935 | 398 | 1139 | 367 | 92.2 | 14 | 3.5 | 1 | 0.3 | 8 | 2.0 | 8 | 2.0 |
| **Age group**  **(in years)** | 0 - 17 | 15135 | 88 | 581 | 86 | 97.7 | 1 | 1.1 | 0 | 0.0 | 1 | 1.1 | 0 | 0.0 |
|  | 18 - 44 | 32507 | 372 | 1144 | 356 | 95.7 | 3 | 0.8 | 1 | 0.3 | 3 | 0.8 | 9 | 2.4 |
|  | 45 - 59 | 13114 | 185 | 1411 | 167 | 90.3 | 9 | 4.9 | 1 | 0.5 | 1 | 0.5 | 7 | 3.8 |
|  | ≥60 | 8679 | 118 | 1360 | 92 | 78.0 | 11 | 9.3 | 0 | 0.0 | 11 | 9.3 | 4 | 3.4 |

*All events that has occurred in the cohort is defined here, before the exclusions based on vaccine type and dates were applied.

| **Supplementary Table 5. COVID-19 incidence and its outcomes among the individuals ≥45 years in the  COVID-19 vaccine effectiveness cohort study, Chennai, 2021 (N=21,793)** | | | | | | | | | | | | | | |
| --- | --- | --- | --- | --- | --- | --- | --- | --- | --- | --- | --- | --- | --- | --- |
|  |  | **Total** | **COVID-19 positive** | **Incidence per 100,000 population** | **Recovered without oxygen support** | | **Recovered with Oxygen Support** | | **Recovered with ventilator support** | | **Died** | | **No Information** | |
| **Characteristic** |  |  | **N** |  | **n** | **%** | **n** | **%** | **n** | **%** | **n** | **%** | **n** | **%** |
| **Overall** |  | 21,793 | 303 | 1390 | 259 | 85.5 | 20 | 6.6 | 1 | 0.3 | 12 | 4.0 | 11 | 3.6 |
| **Gender** | Male | 10916 | 145 | 1328 | 123 | 84.8 | 8 | 5.5 | 0 | 0.0 | 7 | 4.8 | 7 | 4.8 |
|  | Female / TG | 10877 | 158 | 1453 | 136 | 86.1 | 12 | 7.6 | 1 | 0.6 | 5 | 3.2 | 4 | 2.5 |
| **Age Group  (in years)** | 45 - 59 | 13114 | 185 | 1411 | 167 | 90.3 | 9 | 4.9 | 1 | 0.5 | 1 | 0.5 | 7 | 3.8 |
|  | ≥60 | 8679 | 118 | 1360 | 92 | 78.0 | 11 | 9.3 | 0 | 0.0 | 11 | 9.3 | 4 | 3.4 |

| **Supplementary Table 6. Effectiveness of one dose of Covishield vaccine against COVID-19 infection among  individuals ≥45 years, Chennai, India (N=11,668)** | | | | | | | |
| --- | --- | --- | --- | --- | --- | --- | --- |
| **Characteristics & its category** | | **Vaccination status** | **Total** | **COVID-19 positive** | | **RR  (95% CI)** | **VE (95% CI)** |
|  |  |  | **N** | **n** | **%** |  |  |
| **Overall** | **≥45 years** | Received one dose | 4014 | 40 | 1.0 | 0.713 (0.497 - 1.023) | 28.7 (-2.3 - 50.3) |
|  |  | Unvaccinated | 7654 | 107 | 1.4 |  |  |
| **Age Group** | **45-59 years** | Received one dose | 2798 | 28 | 1.0 | 0.694 (0.448 - 1.074) | 30.6 (-7.4 - 55.2) |
|  |  | Unvaccinated | 4713 | 68 | 1.4 |  |  |
|  | **≥60 years** | Received one dose | 1216 | 12 | 1.0 | 0.744 (0.391 - 1.416) | 25.6 (-41.6 - 60.9) |
|  |  | Unvaccinated | 2941 | 39 | 1.3 |  |  |
| **Gender** | **Male** | Received one dose | 2012 | 20 | 1.0 | 0.719 (0.430 - 1.202) | 28.1 (-20.2 - 57.0) |
|  |  | Unvaccinated | 3687 | 51 | 1.4 |  |  |
|  | **Female / TG** | Received one dose | 2002 | 20 | 1.0 | 0.708 (0.426 - 1.176) | 29.2 (-17.6 -57.4) |
|  |  | Unvaccinated | 3967 | 56 | 1.4 |  |  |
| **Comorbidity** | **HT/DM** | Received one dose | 1003 | 12 | 1.2 | 0.910 (0.457 - 1.812) | 9.0 (-81.2 - 54.3) |
|  |  | Unvaccinated | 1826 | 24 | 1.3 |  |  |
|  | **Non-HT/DM** | Received one dose | 3008 | 28 | 0.9 | 0.654 (0.427 - 1.001) | 34.6 (-0.081 - 57.3) |
|  |  | Unvaccinated | 5828 | 83 | 1.4 |  |  |
| **Person per room** | **< 3** | Received one dose | 2619 | 21 | 0.8 | 0.506 (0.312 -0.821) | 49.4 (17.9 - 68.8) |
|  |  | Unvaccinated | 4610 | 73 | 1.6 |  |  |
|  | **>= 3** | Received one dose | 1395 | 19 | 1.4 | 1.219 (0.698 - 2.13) | 18.0 (-43.3 - 53.1) |
|  |  | Unvaccinated | 3044 | 34 | 1.1 |  |  |
| **Residential Area** | **Slum** | Received one dose | 597 | 0 | 0.0 | 0.081 (0.005 - 1.337) | 92.0 (-33.7 - 99.5) |
|  |  | Unvaccinated | 1683 | 18 | 1.1 |  |  |
|  | **Non-Slum** | Received one dose | 3417 | 40 | 1.2 | 0.785 (0.542 - 1.138) | 21.5 (-13.8 - 45.8) |
|  |  | Unvaccinated | 5971 | 89 | 1.5 |  |  |
| *VE - Vaccine effectiveness \| C.I - Confidence interval \| RR - Relative risk* | | | | | | | |

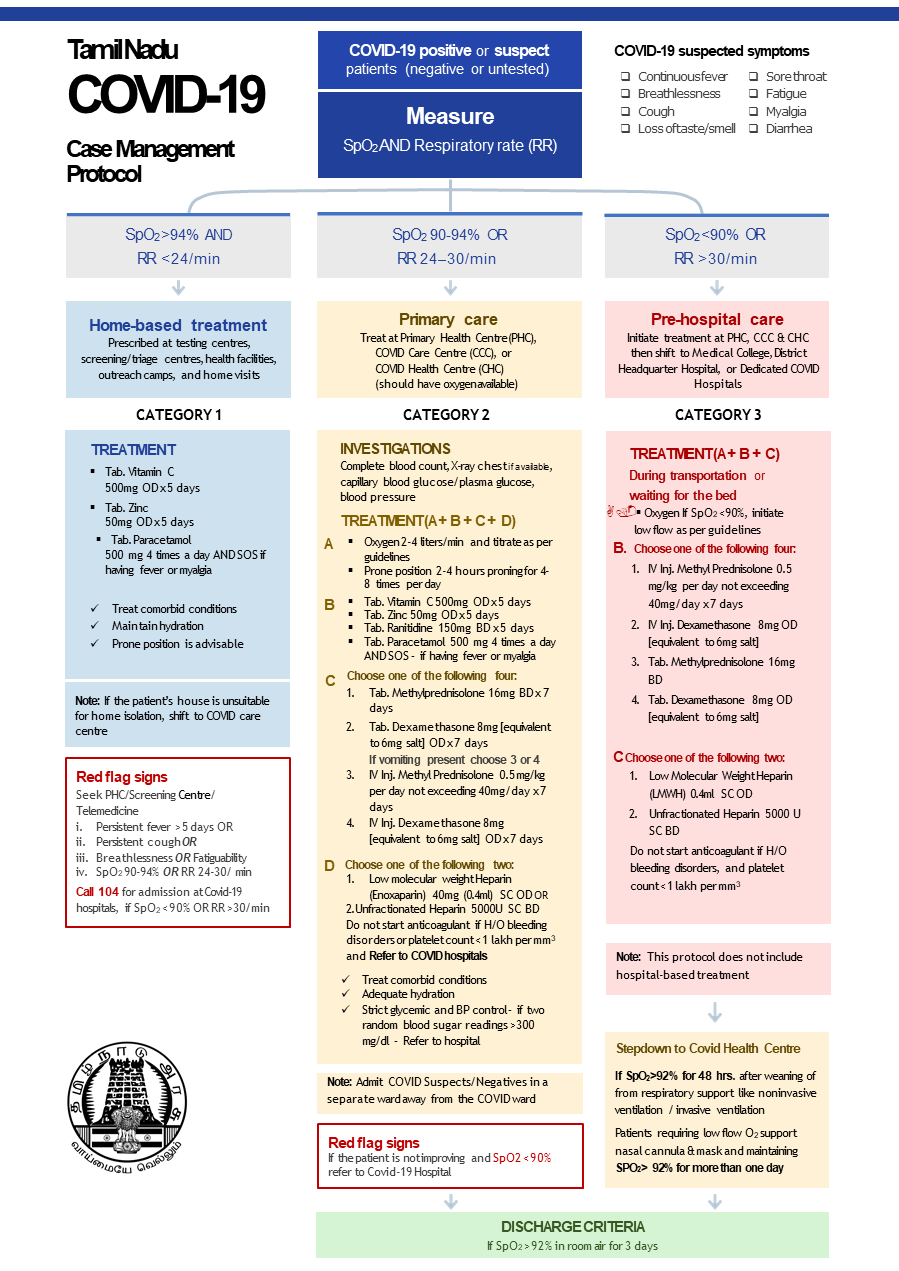

Supplementary Figure 1: Tamil Nadu COVID-19 Case Management Protocol

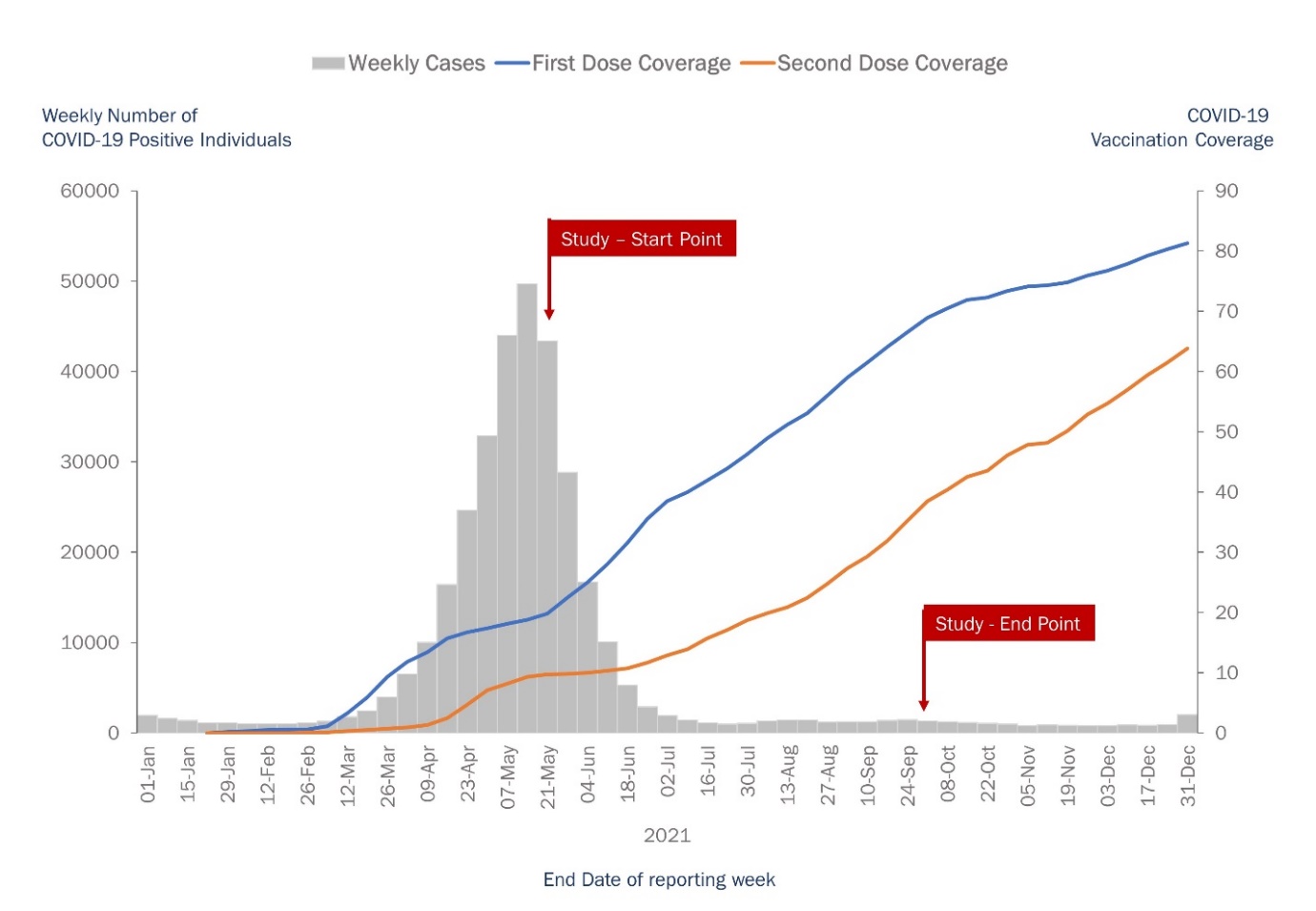

Supplementary Figure 2: Vaccine coverage among the eligible population (18+) and the total number of reported COVID-19 cases by week, Chennai, Tamil Nadu, India, January – December 2021

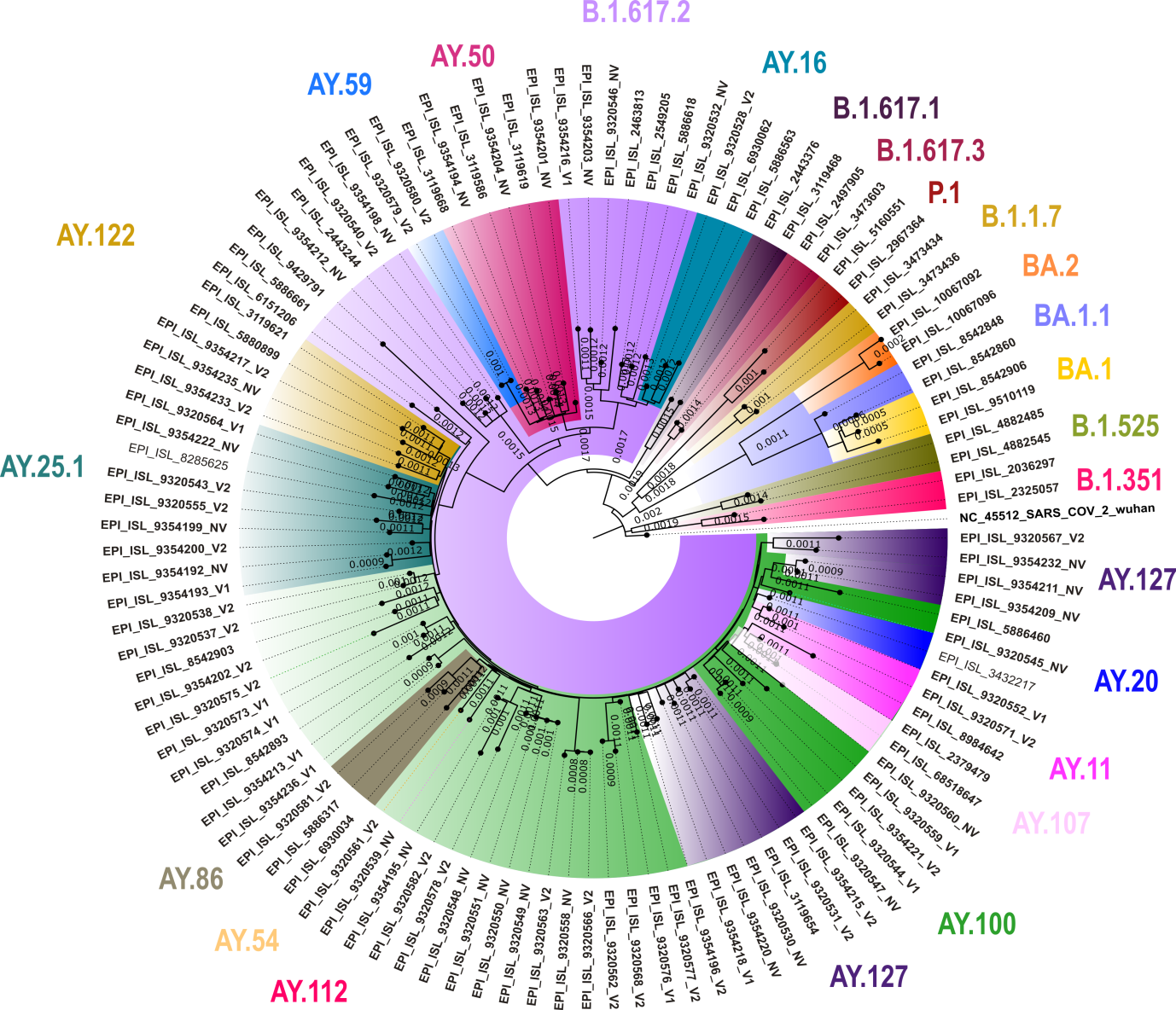

Supplementary Figure 3: Neighbour-joining tree of the SARS-CoV-2 sequences retrieved from the clinical samples collected from the 45+ age group individuals who were positive for SARS-CoV-2 in COVID-19 Vaccine effectiveness Study, Chennai, India, 2021. We generated the tree using MEGA version 10 with Tamura-3-parameter model with a gamma distribution as the rate parameter having bootstrap replication of 1000 cycles. The coloured text represents the lineages. We visualized the generated tree using FigTree v1.4.4.

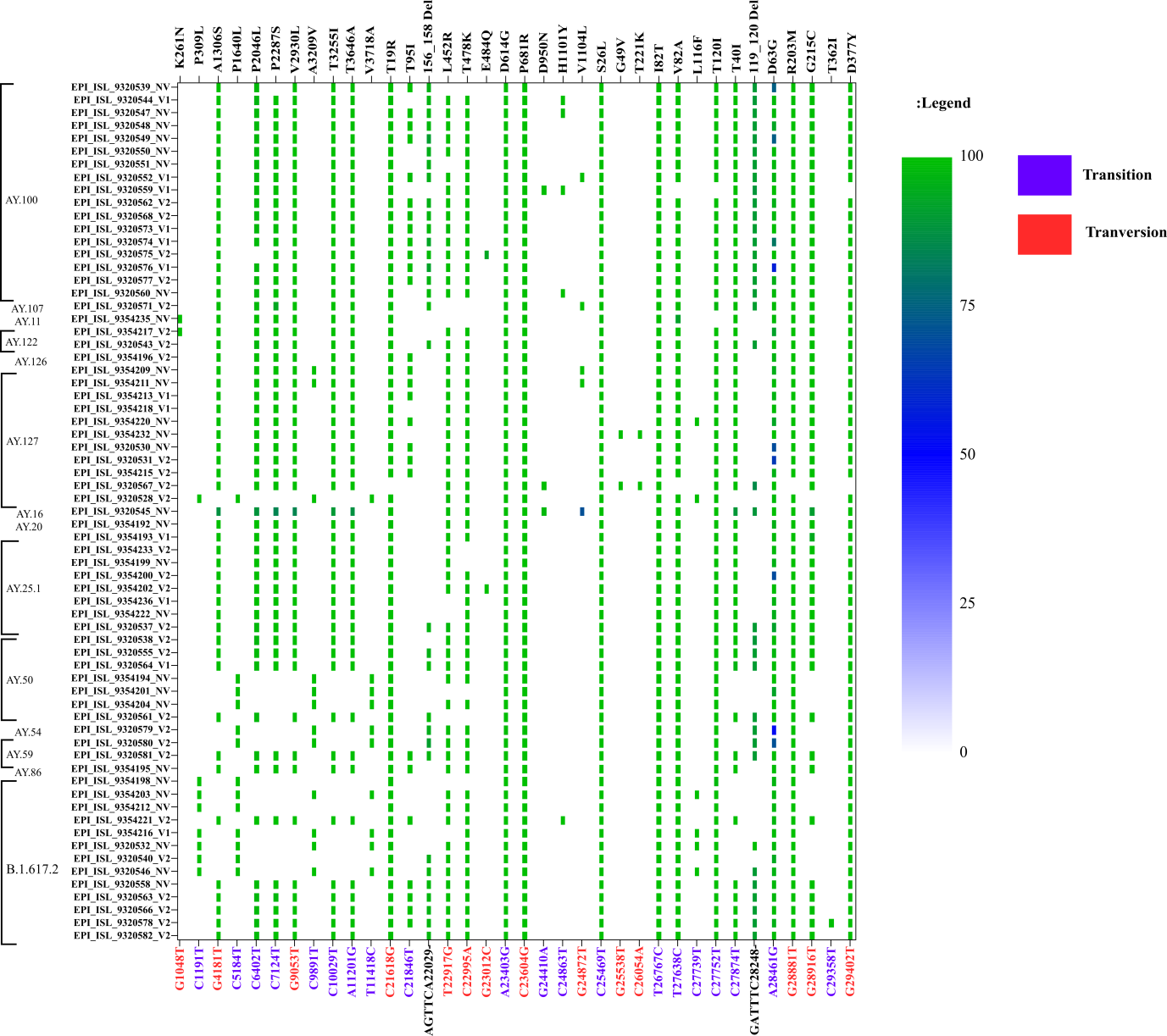

Supplementary Figure 4: Single nucleotide variations in the SARS CoV-2 sequences retrieved from the clinical samples collected from the 45+ age group individuals who were positive for SARS-CoV-2 in COVID-19 Vaccine effectiveness Study, Chennai, India, 2021.

STROBE Statement

|  | Item No | Recommendation | Page No |
| --- | --- | --- | --- |
| **Title and abstract** | 1 | (*a*) Indicate the study’s design with a commonly used term in the title or the abstract | 3 |
|  |  | (*b*) Provide in the abstract an informative and balanced summary of what was done and what was found | 3 |
| Introduction | | | |
| Background/rationale | 2 | Explain the scientific background and rationale for the investigation being reported | 4 |
| Objectives | 3 | State specific objectives, including any prespecified hypotheses | 5 |
| Methods | | | |
| Study design | 4 | Present key elements of study design early in the paper | 6 |
| Setting | 5 | Describe the setting, locations, and relevant dates, including periods of recruitment, exposure, follow-up, and data collection | 6 |
| Participants | 6 | (*a*) Give the eligibility criteria, and the sources and methods of selection of participants | 7 |
| Variables | 7 | Clearly define all outcomes, exposures, predictors, potential confounders, and effect modifiers. Give diagnostic criteria, if applicable | 8-9 |
| Data sources/ measurement | 8* | For each variable of interest, give sources of data and details of methods of assessment (measurement). Describe comparability of assessment methods if there is more than one group | 8-9 |
| Bias | 9 | Describe any efforts to address potential sources of bias | 8-9 |
| Study size | 10 | Explain how the study size was arrived at | 7-8 |
| Quantitative variables | 11 | Explain how quantitative variables were handled in the analyses. If applicable, describe which groupings were chosen and why | 10 |
| Statistical methods | 12 | (*a*) Describe all statistical methods, including those used to control for confounding | 10-11 |
|  |  | (*b*) Describe any methods used to examine subgroups and interactions | 11 |
|  |  | (*c*) Explain how missing data were addressed | NA |
|  |  | (*d*) If applicable, describe analytical methods taking account of sampling strategy | NA |
|  |  | (*e*) Describe any sensitivity analyses | NA |
| Results | | | |
| Participants | 13* | (a) Report numbers of individuals at each stage of study—eg numbers potentially eligible, examined for eligibility, confirmed eligible, included in the study, completing follow-up, and analysed | 12 |
|  |  | (b) Give reasons for non-participation at each stage | 12 |
|  |  | (c) Consider use of a flow diagram | 12 |
| Descriptive data | 14* | (a) Give characteristics of study participants (eg demographic, clinical, social) and information on exposures and potential confounders | 12 |
|  |  | (b) Indicate number of participants with missing data for each variable of interest | NA |
| Outcome data | 15* | Report numbers of outcome events or summary measures | 12-13 |
| Main results | 16 | (*a*) Give unadjusted estimates and, if applicable, confounder-adjusted estimates and their precision (eg, 95% confidence interval). Make clear which confounders were adjusted for and why they were included | 13 |
|  |  | (*b*) Report category boundaries when continuous variables were categorized | 12-13 |
|  |  | (*c*) If relevant, consider translating estimates of relative risk into absolute risk for a meaningful time period | NA |
| Other analyses | 17 | Report other analyses done—eg analyses of subgroups and interactions, and sensitivity analyses | 13-14 |
| Discussion | | | |
| Key results | 18 | Summarise key results with reference to study objectives | 15-16 |
| Limitations | 19 | Discuss limitations of the study, taking into account sources of potential bias or imprecision. Discuss both direction and magnitude of any potential bias | 17 |
| Interpretation | 20 | Give a cautious overall interpretation of results considering objectives, limitations, multiplicity of analyses, results from similar studies, and other relevant evidence | 17-18 |
| Generalisability | 21 | Discuss the generalisability (external validity) of the study results | 17 |
| Other information | | | |
| Funding | 22 | Give the source of funding and the role of the funders for the present study and, if applicable, for the original study on which the present article is based | 19 |

*Give information separately for exposed and unexposed groups.
